# Epidemiological, clinical, and laboratory findings for patients of different age groups with confirmed coronavirus disease 2019 (COVID-19) in a hospital in Saudi Arabia

**DOI:** 10.1101/2020.10.21.20217083

**Authors:** Mutasim E. Ibrahim, Obaid S. AL-Aklobi, Mosleh M. Abomughaid, Mushabab A. Al-Ghamdi

**Affiliations:** Department of Basic Medical Sciences (Microbiology Unit), College of Medicine, University of Bisha, Bisha, Saudi Arabia; Director of Infection Prevention and Control, Bisha Health Directorate, Ministry of Health, Bisha, Saudi Arabia; Department of Medical Laboratory Sciences, College of Applied Medical Sciences, University of Bisha, Bisha, Saudi Arabia; Department of Internal Medicine, College of Medicine, University of Bisha, Bisha, Saudi Arabia; Internal Medicine Department, King Abdullah Hospital, Ministry of Health, Bisha, Saudi Arabia

**Keywords:** COVID-19, epidemiology, clinical characteristics, laboratory findings, Saudi Arabia

## Abstract

**Background:** Although the coronavirus disease 2019 (COVID-19) pandemic continues to rage worldwide, clinical and laboratory studies of this disease have been limited in many countries. We investigated the epidemiologic, clinical, and laboratory findings of COVID-19 infected patients to identify the effective indicators correlated with the disease.

**Methods:** A retrospective study was conducted at King Abdullah Hospital in Bisha Province, Saudi Arabia, from March 20 to June 30, 2020. Patients of different age groups were confirmed as having COVID-19 infection using a real-time polymerase chain reaction. The demographic, clinical, and laboratory data of the patients were statistically analyzed.

**Results:** Of the 137 patients, 88 were male and 49 were female, with a mean age of 49.3 years (SD±18.4). The patients were elderly (n=29), adults (n=103), and children (n=5). Of these, 54 (39.4%) had comorbidities, 24% were admitted to the intensive care unit (ICU), and 12 (8.8%) died. On admission, the main clinical manifestations were fever (82.5%), cough (63.5%), shortness of breath (24.8%), chest pain (19.7%), and fatigue (18.2%).

In all patients, increased neutrophils and decreased lymphocytes were observed. Patients’ lactate dehydrogenase (LDH) was elevated. C-reactive protein (CRP) was elevated in 46.7%, D-dimer in 41.6%, and the erythrocyte sedimentation rate (ESR) in 39.4% of patients. The elderly showed higher neutrophil (p=0.003) and lower lymphocyte (p=0.001) counts than adults and children. Glucose, creatine kinase-MB, LDH, bilirubin, D-dimer, and ESR were significantly higher in the elderly than in the adults. The COVID-19 death group had a higher leucocyte count (p = 0.043), and higher urea (p=0.025) and potassium (p=0.026) than the recovered group but had a lower hemoglobin concentration (p=0.018). A significant association was determined between COVID-19 death (χ2(1)=17.751, p<0.001), and the presence of cardiovascular disease (χ2(1)=17.049, p<0.001), hypertension (χ2(1)=7.659, p=0.006), renal failure (χ2(1)=4.172, p<0.04), old age (t(135) = 4.747, p <0.001), and ICU admission (χ2(1) = 17.751 (1), p<0.001).

**Conclusions:** The common symptoms found in this study could be useful for identifying potential COVID-19 patients. Investigating some of the laboratory and clinical parameters could help assess the disease progression, risk of mortality, and follow up patients who could progress to a fatal condition.

## Introduction

The novel coronavirus designated as 2019-nCoV is a human pathogen discovered in China at the end of 2019 during an outbreak of endemic pneumonia of unknown etiology [1,2].The World Health Organization (WHO) named this new coronavirus SARS-CoV-2, and the disease coronavirus disease 2019 (COVID-19) [2]. The disease continues to spread in more than 200 countries worldwide, with a high rate of infection and mortality, creating a significant dilemma for global health [1,3].

The COVID-19 infection is represented by a spectrum of clinical severity. From no symptoms or mild upper respiratory tract symptoms [4–6], the disease can develop to severe pneumonia characterized by fever, cough, fatigue, dyspnea, bilateral pulmonary infiltrates, and acute respiratory injury[4]. The clinical characteristics and severity of COVID-19 have been described as similar to other coronaviruses, such as Middle East respiratory syndrome (MERS) and severe acute respiratory syndrome (SARS) [3]. Patients with severe illness can develop acute respiratory distress syndrome (ARDS), which requires extended care in an intensive care unit [6,7] Recent studies have confirmed that critically ill patients develop severe complications such as septic shock, severe metabolic acidosis, coagulopathy, thrombocytopenia, arrhythmias, and multiple organ dysfunction syndrome[5,8,9].

In Saudi Arabia, the first confirmed case of COVID-19 was recorded on March 2, 2020, and the number of discovered cases is increasing [10]. By September 27, 2020, the Saudi Ministry of Health had reported a cumulative total of 332,790 cases and 4,655 deaths confirmed in Saudi Arabia [11]. Such a large number of cases highlights the importance of analyzing the characteristics of patients confirmed to have the disease[3]. Recent studies have noted that the clinical and laboratory features of the disease are variable between populations because of their distinct demographic features, comorbidities, and immune system responses [8,12]. However, testing the epidemiological, clinical, and laboratory elements correlated with COVID-19 infections is important for determining high-risk groups, mapping the disease, and designing appropriate management [8]. To date, multiple studies have described the disease among various population groups, but these have been mainly in China [4,7,12,13]. Published data concerning the epidemiological, clinical, and laboratory findings of the disease in Saudi Arabia are limited [3,8,14]. This study investigated the potential role of the epidemiological, clinical, and laboratory findings of COVID-19 infected patients of different ages to identify the effective indicators associated with the disease.

## Methodology

### Study design and setting

A retrospective single-center study was conducted at King Abdullah Hospital (KAH), Bisha Province, Saudi Arabia. This hospital is a referral hospital with nearly 400 beds. It covers various specialties, thus serving various patient groups and most of the population in different areas in Bisha Province and surrounding villages[15]. KAH is the only hospital that admits COVID-19 patients in Bisha Province and the surrounding rural areas.

### Patients and procedures

Patients having COVID-19 infection who were admitted to KAH between March 20 and June 30, 2020 were enrolled in this study. We categorized patients into three groups, namely children (1 to 14 years), adults (15 to 64 years), and elderly (65 years or older). Patients who were less than 1 year old were excluded from the study due to insufficient clinical and laboratory data. All patients were confirmed as having the virus by positive SARS-CoV-2 nucleic acid results using a real-time reverse transcriptase-polymerase chain reaction (RT-PCR) assay on nasopharyngeal swab specimens. The RT-PCR test for COVID-19 was carried out at the Aseer Regional Hospital, Aseer region, Saudi Arabia, and the test results were available online within 24 h.

### Collection of clinical and laboratory data

Data on the demographic characteristics, clinical manifestations, and laboratory findings were obtained from patients’ medical records and local health authorities, and were entered into data collection form on admission. We also collected laboratory test results, including results for complete blood count, glucose levels, coagulation tests, D-dimer levels, erythrocyte sedimentation rate (ESR), renal and liver function tests, electrolytes, C-reactive protein (CRP), lactate dehydrogenase (LDH), creatine kinase (CK), and CK-MB.

### Ethical clearance

This study was reviewed and approved by the Research Ethics Local Committee at the College of Medicine, University of Bisha (UBCOM/ H-06-BH-087 (05/03)). The data were collected in an anonymous format, without violating the personal privacy of the patients. The study was carried out as part of the response by the University of Bisha to investigate the COVID-19 outbreak.

### Statistical analysis

Data were entered and analyzed using the software Statistical Package for Social Sciences (SPSS version 22) (Armonk, NY: IBM Corp.). Descriptive statistics were calculated to present the baseline demographic data and clinical symptoms in terms of proportion and frequency. Normally distributed continuous data were presented as mean ± standard deviation (SD), and were compared using independent group t-tests. Non-normal distributed continuous variables were examined by the Mann–Whitney U test and expressed as the median and interquartile ranges (IQR). Non-parametric Kruskal–Wallis rank sum was applied for pairwise and multiple comparisons between the groups, and was presented as the median and IQR. Categorical variables were compared using the chi-squared test and displayed as numbers and proportions. All p-values of <0.05 were considered statistically significant.

## Results

One hundred thirty-seven confirmed cases of COVID-19 were admitted to the King Abdullah Hospital during the period from March 18, 2020, to June 30, 2020. The average length of a hospital stay was four days (IQR, 3–7 days). The total number of admitted cases per month was six in March and four in April, which increased 31 in May and 96 in June. Of the137 patients, 88 (64.2%) were male and 49 (35.8%) were female, with a mean age of 49.3 years (SD ±18.4). Most of the patients were adults (75.2%), followed by the elderly (21.2%), and children (3.6%). Most of the patients (73.7%) were Saudi. One hundred and seven patients (78.1%) had direct contact with infected individuals, and 30 (21.9%) had a history of travel outside the country.

### Clinical presentation

Table 1 outlines the main clinical manifestations associated with the disease. On admission, the most common clinical manifestation was fever (82.5%), followed by cough (63.5%), shortness of breath (24.8%), chest pain (19.7%), and fatigue (18.2). Fifty-four (39.4%) of the patients had chronic disease, including diabetes mellitus, renal failure, hypertension, liver disease, cardiovascular disease, and respiratory disease. Thirty-four (24%) of the cases were admitted to the ICU. The mortality rate of the admitted cases was 8.8% (12/137).

**Table 1:** Baseline characteristic of patients with COVID-19 infection.

| **Characteristic** | **n(%)** |
| --- | --- |
| Age group |  |
| 1 to 15years old | 5 (3.6) |
| 16 to 64 | 103 (75.2) |
| 65 and above | 29 (21.2) |
| Gender |  |
| Male | 88 (64.2) |
| Female | 49 (35.8) |
| Nationality |  |
| Saudi | 101 (73.7) |
| Non-Saudi | 36 (26.3) |
| Symptoms |  |
| Fever | 113 (82.5) |
| Cough | 87(63.5) |
| Shortness of breath | 34 (24.8) |
| Chest pain | 27(19.7) |
| Fatigue | 25 (18.2) |
| Headache | 22 (16.1) |
| Sore throat | 13 (9.5) |
| Nausea and vomiting | 10 (7.3) |
| Abdominal pain | 8 (5.8) |
| Congested nose | 6 (4.4) |
| Diarrhea | 5 (3.6) |
| Muscle main | 4 (2.9) |
| Comorbidity | 54 (39.4) |
| Diabetes mellitus | 37 (27.0) |
| Renal failure | 19 (13.9) |
| Hypertension | 14 (10.2) |
| Liver diseases | 5 (3.6) |
| Cardiovascular diseases | 5 (3.6) |
| Respiratory diseases | 2 (1.5) |
| Mortality rate | 12 (8.8) |
| No. of cases transferred to intensive care unit | 34 (24.8) |

### Laboratory results

The results of the laboratory parameters obtained at the time of patient admission are displayed in Table 2. The complete blood count results of all patients were recorded as average values, and showed slightly increased neutrophil and decreased lymphocyte counts. Overall, the average blood biochemistry parameters were normal, but elevated levels were detected for LDH (276 UL, normal values = 81–234). Regarding the inflammatory biomarkers, CRP was increased in 46.7% of the patients; D-dimer was elevated in 41.6% of the patients, and ESR was increased in 39.4% of patients.

**Table 2:** Laboratory findings for patients of different age groups with confirmed COVID-19.

| Parameter | Median (IQR) |  |  |  | P value |  |  |  |
| --- | --- | --- | --- | --- | --- | --- | --- | --- |
|  | Total | Children<sup>a</sup> | Adults<sup>b</sup> | Elderly<sup>c</sup> | a/b/c | a/b | a/c | b/c |
| TLC (4-11)x10<sup>9</sup> | 6.6 (4.8-9.1) | 5.9 (4.3-9.8) | 6.3 (4.7-9.1) | 7.6(5.36-10) | 0.347 | 0.942 | 0.576 | 0.146 |
| Neutrophil (50-70) x10<sup>9</sup> | 67.6 (59.75-76.1) | 56 (50-60.9) | 66.5 (59-76.1) | 73.1(67.6-81.8) | 0.003 | 0.031 | 0.006 | 0.011 |
| Lymphocyte (25-40) x10<sup>9</sup> | 23 (16.55-29.6) | 35 (29.4-40) | 23.9 (16.6-30.3) | 19.6(14.15-23.05) | 0.001 | 0.018 | 0.001 | 0.009 |
| Monocyte (1-8) x10<sup>9</sup> | 7.7 (5-9.5) | 8 (7.5-8.9) | 8 (5-9.5) | 6(3.3-10) | 0.174 | 0.787 | 0.113 | 0.084 |
| Eosinophil (0-4) x10<sup>9</sup> | 0.3 (0-1.5) | 0.5 (0.3-1.55) | 0.3 (0-1.3) | 0.2(0-2.3) | 0.613 | 0.317 | 0.397 | 0.933 |
| Basophil (0-1) 10<sup>9</sup> | 0.4 (0.3-0.6) | 0.5 (0.5-0.75) | 0.4 (0.3-0.6) | 0.4(0.3-0.7) | 0.313 | 0.182 | 0.289 | 0.415 |
| Hemoglobin (12-16) g/dl | 13.6 (11.85-15.1) | 12.8 (11.55-13.95) | 13.7 (11.9-15.1) | 12.6(11.6-14.9) | 0.358 | 0.292 | 0.846 | 0.267 |
| Platelets (130-400) x10<sup>9</sup> | 229 (178-302) | 252 (157.5-338.5) | 231 (184-314) | 196 (164-244) | 0.043 | 0.693 | 0.206 | 0.014 |
| PT (9.0-14) second | 11.5 (10.95-12.15) | 11.9 (11.65-12.85) | 11.3 (10.9-12.2) | 11.8(10.8-12) | 0.375 | 0.153 | 0.272 | 0.817 |
| INR | 1 (0.9-1.0) | 1 (0.85-1.25) | 1 (0.9-1.1) | 1(0.9-1.0) | 0.891 | 0.933 | 0.939 | 0.63 |
| Glucose (3.9-11) mmol/L | 6.7 (5.6-10.5) | 5.4 (4-11.75) | 6.2 (5.4-7.9) | 8.7(6.45-15.65) | 0.007 | 0.276 | 0.072 | 0.003 |
| Urea (1.7-8.3) mmol/L | 4.6 (3.35-6.1) | 3.3 (2.8-5.05) | 4.2 (3.3-5.5) | 6.4(4.25-12.35) | <0.001 | 0.331 | 0.027 | <0.001 |
| Creatinine (15-115) umol/L | 83 (67.5-103.5) | 46 (36.8-64) | 77 (67-98) | 103(80-139.5) | <0.001 | 0.002 | 0.001 | 0.001 |
| Albumin (34-50) g/L | 33 (29.4-36) | 30 (28.5-31.5) | 33 (30-37) | 33(28.75-35.75) | 0.173 | 0.08 | 0.18 | 0.415 |
| ALT (0.0-41) U/L | 29 (19.5-49.5) | 15 (7.65-53.5) | 34 (20-51) | 29(20-49) | 0.251 | 0.105 | 0.119 | 0.777 |
| AST (0.0-38) U/L | 34 (20.5- 55.5) | 30.8 (15-102.5) | 30 (18-55) | 41(27-61.5) | 0.21 | 0.884 | 0.576 | 0.076 |
| CKMB (0-25) U/L | 18 (14-24) | 15 (13-77.5) | 17 (14-23) | 23(16.5-29.5) | 0.089 | 0.543 | 0.284 | 0.035 |
| CK (26-192) U/L | 118 (62-244.5) | 198 (72.5-545) | 104 (60-227) | 166 (73-317.5) | 0.248 | 0.273 | 0.808 | 0.168 |
| LDH (81-234) U/L | 276 (174-345.5) | 256 (233.5-380) | 260 (167-342) | 322 (207-448.5) | 0.071 | 0.474 | 0.827 | 0.024 |
| Na<sup>+</sup> (135-145) mmol/L | 136 (132-138) | 139 (138-140) | 136 (133-138) | 134 (131-138.5) | 0.029 | 0.01 | 0.04 | 0.313 |
| K<sup>+</sup> (3.5-5.0) mmol/L | 4 (3.7-4.4) | 4.4 (3.95-4.8) | 3.98 (3.6-4.3) | 4.1(3.74-4.95) | 0.097 | 0.096 | 0.679 | 0.109 |
| Bilirubin (0.0-18.1) umol/L | 6.1 (4.8-10.9) | 6.4 (5.6-9.4) | 6.3 (4.7-9.1) | 9.4(5.1-12.3) | 0.073 | 0.206 | 0.827 | 0.043 |
| *CRP, n(%) |  |  |  |  |  |  |  | 0.216 |
| Increased | 64 (46.7) | 0 (0.0) | 47 (45.6) | 17 (58.6) |  |  |  |  |
| Normal | 73 (53.3) | 5 (100) | 56 (54.4) | 12 (41.4) |  |  |  |  |
| *D-Dimer, n(%) |  |  |  |  |  |  |  | 0.006 |
| Increased | 57 (41.6) | 0 (0.0) | 38 (36.9) | 19 (65.5) |  |  |  |  |
| Normal | 80 (58.4) | 5 (100) | 65 (63.1) | 10 (34.5) |  |  |  |  |
| *ESR, n(%) |  |  |  |  |  |  |  |  |
| Increased | 54 (39.4) | 0 (0.0) | 36 (35) | 18 (62.1) |  |  |  | 0.009 |
| Normal | 83 (60.6) | 5 (100) | 67 (65) | 11 (37.9) |  |  |  |  |

ALT alanine transaminase, AST aspartate aminotransferase, CK creatine kinase, CRP C-reactive protein, ESR erythrocyte sedimentation rate, INR international, IQR interquartile range, K<sup>+</sup> potassium normalized ratio, LDH lactate dehydrogenase, Na<sup>+</sup> sodium, PT prothrombin time, TLC total leucocyte count
\* Chi-square used to compare between elderly and adult groups

### Comparison of laboratory parameters by age group

Table 2 compares the laboratory findings of the three age groups of children, adults, and the elderly. Neutrophil counts were significantly higher in the elderly group (median = 73.1, IQR [67.6–81.8]) than in adults (median = 66.5, IQR [59–76.1]) or children (median = 56, IQR [50– 60.9]) groups. Lymphocytes were significantly higher in the children (median = 35, IQR [29.4– 40]) than in the adult (median = 23.9, IQR [16.6–30.3]) or elderly groups (median = 19.6, IQR [14.15–23.05]). Platelet counts were significantly lower in the elderly than in adult patients (p = 0.014). There were no significant differences in other blood count parameters between the three age groups.

In terms of blood biochemistry results, glucose, CK-MB, LDH, and bilirubin were significantly higher in the elderly than in the adults (Table 2). Significant differences (p <0.001) in creatinine levels were determined between the three groups, with the highest level in the elderly group (median = 103, IQR [80–139.5]), followed by adults (median = 77, [IQR, 67–98]) and children (median = 46, IQR [36.8–64]). There were no significant differences in alanine transaminase (ALT), aspartate aminotransferase (AST), albumin, CK, and potassium levels among the three groups (Table 2).

As shown in Table 2, the number of patients with increased D-dimer and ESR was significantly higher in the elderly group compared with the adult group ([D-dimer: 65.6% vs. 36.9; p = 0.006], [ESR: 62.1% vs. 35%; p = 0.009). The number of patients with increased CRP levels did not significantly differ between the elderly and adult groups (58.6% vs. 45.6%; p = 0.216).

### Factors associated with COVID-19 death

In Table 3, the analysis of several factors in relation to COVID-19 death is shown. There was a statistically significant association between COVID-19 death and ICU admission (χ2(1) = 17.751, p < 0.001), cardiovascular disease (χ2(1) = 17.049, p < 0.001), hypertension (χ2(1) = 7.659, p = 0.006), and renal failure (χ2(1) = 4.172, p = <0.04). Moreover, COVID-19 death was significantly increased (t(135) = 4.747, p <0.001) in the elderly group (mean = 71.7, SD [17.0]) compared with the adult and children groups (mean = 47.2, SD [17.1]). However, there was no significant effect for sex (χ2(1) = 2.915, p < 0.088), although men (n = 88) were more likely to be infected by the disease than women (n = 49).

**Table 3:** Factors associated with COVID-19 death.

| Variable | Total number | Improved | No of death (%) | df | P value |
| --- | --- | --- | --- | --- | --- |
| Gender |  |  |  | 2.915 (1) | 0.088 |
| Male | 88 | 83 | 5 (41.7) |  |  |
| Female | 49 | 42 | 7 (58.3) |  |  |
| Nationality |  |  |  | 2.186 (1) | 0.139 |
| Saudi | 101 | 90 | 11 (10.9) |  |  |
| Non-Saudi | 36 | 35 | 1 (2.8) |  |  |
| ICU admission |  |  |  | 17.751 (1) | <0.001 |
| Yes | 34 | 25 | 9 (26.5) |  |  |
| No | 103 | 100 | 3 (2.9) |  |  |
| Hypertension |  |  |  |  | 0.006 |
| Yes | 14 | 10 | 4 (28.6) | 7.659 (1) |  |
| No | 123 | 115 | 8 (6.5) |  |  |
| Renal failure |  |  |  | 4.172 (1) | 0.041 |
| Yes | 19 | 15 | 4 (21.1) |  |  |
| No | 118 | 110 | 8 (6.8) |  |  |
| Diabetes mellitus |  |  |  | 0.205 (1) | 0.650 |
| Yes | 38 | 34 | 4 (10.5) |  |  |
| No | 99 | 91 | 8 (8.1) |  |  |
| Liver diseases |  |  |  | 0.498 (1) | 0.480 |
| Yes | 5 | 5 | 0 |  |  |
| No | 132 | 120 | 12 (100) |  |  |
| Cardiovascular disease |  |  |  | 17.049 (1) | <0.001 |
| Yes | 5 |  | 23 (60) |  |  |
| No | 132 | 123 | 9 (6.8) |  |  |
| Respiratory diseases |  |  |  | 0.135 (1) | 0.659 |
| Yes | 2 |  | 0 |  |  |
| No | 135 |  | 12 (100) |  |  |
| Length of hospital stay, median (IQR) |  | 4 (3-7) | 4.5 (3-7) |  | 0.803 |
| Age, (Mean±SD) | 49.3±18.6 | (47.2±17.1) | (71.7±17.0) | 4.747 (135) | <0.001 |

### Laboratory findings associated with COVID-19 death

Compared with the recovered group, the COVID-19 death group had significantly higher total leucocyte count (median = 8.78, IQR [5.73–13.50] vs. (median = 6.39, IQR [4.7–8.45]; p = 0.043), urea (median = 7.05, IQR [3.90–12.93] vs. median = 4.5, IQR [3.3–5.7]; p = 0.025), and potassium (median = 4.5, IQR [3.96–4.90] vs. median = 3.98, IQR [3.68–4.3]; p = 0.026). In contrast, decreased levels of hemoglobin (median = 11.75, IQR [10.90–13.20] vs. (median = 13.7, IQR [11.9–15.15]; p = 0.018) were reported in the COVID-19 death group compared with the recovered group. There were no statistical differences in the results of other tested laboratory parameters between the recovered and death groups (Table 4).

**Table 4:** Laboratory investigation associated with COVID-19 death.

| Parameter | Improved | Dead | P value |
| --- | --- | --- | --- |
| **Hematology and coagulation, median (IQR)** |  |  |  |
| TLC | 6.39 (4.7-8.45) | 8.78 (5.73-13.50) | 0.034 |
| Neutrophil | 67.1(59.45-74.55) | 74.45 (68.15-81.43) | 0.066 |
| Lymphocyte | 23.1(16.6-29.75) | 18.4 (14.80-21.63) | 0.068 |
| Monocyte | 7.8 (5.35-9.5) | 5.25 (3.15-9.13) | 0.082 |
| Eosinophil | 0.3(0-1.5) | 0.2 (0.00-0.80) | 0.423 |
| Basophil | 0.5 (0.3-0.6) | 0.2 (0.13-0.53) | 0.017 |
| Hemoglobin | 13.7 (11.9-15.15) | 11.75 (10.90-13.20) | 0.018 |
| Platelets count | 229 (178-307) | 198.5 (151.00-289.50) | 0.411 |
| PT | 11.5 (11-12.2) | 11.35 (10.58-11.98) | 0.346 |
| INR | 1(0.9-1.05) | 0.95 (0.90-1.00) | 0.697 |
| **Blood biochemistry, median (IQR)** |  |  |  |
| Glucose | 6.4 (5.6-9.2) | 9.75 (5.68-15.78) | 0.166 |
| Urea | 4.5 (3.3-5.7) | 7.05 (3.90-12.93) | 0.025 |
| Creatinine | 80 (67.5-102) | 100 (69.50-135.50) | 0.205 |
| Albumin | 33 (29.9-36) | 32.3 (24.50-38.60) | 0.55 |
| ALT | 32 (20-51) | 27 (14.50-38.25) | 0.286 |
| AST | 33 (20-55.5) | 41(29.25-60.75) | 0.281 |
| CK-MB | 7 (14-24) | 20.5 (1.18-32.00) | 0.228 |
| CK | 118 (62- | 155 (52.75-240.75) | 0.837 |
| LDH | 273 (174-345.5) | 321(161.50-357.00) | 0.933 |
| Na | 136 (132-138) | 135.5 (132.50-139.75) | 0.708 |
| K | 3.98 (3.68-4.3) | 4.5 (3.96-4.90) | 0.026 |
| Bilirubin | 6.1 (-10.9) | 5.65 (3.60-11.58) | 0.420 |
| **Inflammatory biomarkers, n(%)** |  |  |  |
| CRP |  |  | 0.294 |
| Normal | 68 | 5/12 (41.7) |  |
| Increased | 57 | 7/12 (58.3) |  |
| D-dimer |  |  | 0.065 |
| Normal | 76 | 4/12 (33.3) |  |
| Increased | 49 | 8/12 (6.7) |  |
| ESR |  |  | 0.160 |
| Normal | 78 | 5/12 (41.7) |  |
| Increased | 47 | 7/12 (58.3) |  |

ALT alanine transaminase, AST aspartate aminotransferase, CK creatine kinase, CRP C-reactive protein, ESR erythrocyte sedimentation rate, INR international, K<sup>+</sup> potassium normalized ratio, IQR interquartile range, LDH lactate dehydrogenase, Na<sup>+</sup> sodium, PT prothrombin time, TLC total leucocyte count

## Discussion

In this study, we described the epidemiology, clinical characteristics, and laboratory parameters of 137 confirmed patients with COVID-19 in Bisha, Saudi Arabia. We found that COVID-19 frequently infected the middle-aged group. This is in agreement with previous reports that showed the middle-aged, from 40 to 60 years, were the most commonly infected group [16–19]. However, the virus can infect individuals of all age groups[17,18] The study reported a greater number of men than women as having the infection, which is similar to the findings of other studies [19–21]. A study conducted among 56 elderly patients found that more men than women were infected by 2019-nCoV [13]. A single□arm meta□analysis showed that the men made up a larger percentage of the gender distribution of COVID□19 patients at 60% (95% CI [0.54, 0.65])[20]. Previous studies have documented more men than women to be infected with other coronavirus infections, such as in SARS-CoV[22] and MERS-CoV[23]. The decrease in women’s susceptibility to coronavirus infections could be attributed to sex hormones and an X chromosome, which play an essential role in innate and adaptive immunity[19,20]

Our findings revealed that fever, cough, and shortness of breath were the most frequent symptoms in COVID-19 patients, which is in agreement with many previous studies [13,24–26]. Research evidence suggests that the disease’s complete clinical characteristics are nonspecific and are not yet clear, as the associated symptoms range from mild to severe, with several cases resulting in death[11,27] However, it is unclear whether the viral load affects the clinical presentation [18]. It is well known that the clinical manifestations in symptomatic patients begin to present with mild symptoms in less than 1 week, and that patients tend to recover within 1 week[14,27] Some patients may progress to severe disease with dyspnea, which may be accompanied by pneumonia in the second or third weeks of the disease [9,27,28]. Severe cases have been found to develop to progressive respiratory failure due to alveolar damage from the virus, leading to death [14,27].

Regarding the hematological parameters, an estimation of white blood counts is essential to predicting the outcomes of COVID-19 infection[8]. We found normal leucocyte counts in our results, with a decreased level of lymphocytes and an upper limit of neutrophil counts. Similarly, Jin et al. showed that in the early stage of the disease, the total number of leucocytes in peripheral blood was normal or decreased, and the lymphocyte count was decreased [26]. Recent reports have also found increased neutrophil counts and decreased lymphocyte counts associated with COVID-19 [29,30] Lymphocytopenia could explain the effect of SARS-CoV-2 on T lymphocytes, mainly on CD4+ T cells and CD8+ T cells, resulting in a decrease in lymphocyte numbers and IFN-γ production[19,30]. It is well known that SARS-CoV-2 spreads through the respiratory mucosa and invades other cells, induces a series of immune responses, and causes changes in peripheral white blood cells, including lymphocytes [19]. Studies suggest that a substantial decrease in the total number of lymphocytes indicates that coronaviruses disrupt many immune cells and suppress cellular immune function [31,32].

In comparing hematological findings, we detected significantly higher neutrophil levels among elderly patients compared with adult or child patients. Similar findings have been reported by others, where older patients with COVID-19 infections have shown high neutrophil counts and decreased lymphocytes[13,29] The elevated neutrophil counts in this study could be explained by the fact that elderly 2019-nCoV infected patients are more susceptible to bacterial infection[13,25]. This study also found that lymphocyte counts were significantly lower in elderly than in adult and child patients. In a retrospective study compared the clinical features of COVID-19 in elderly patients with young and middle-aged patients, Liu et al. found that the proportion of lymphocytes in the elderly group was significantly lower than that in the young and middle-aged group (p < 0.001)[13]. The changes in the muscles that aid respiration, and the anatomical changes in the lungs associated with an older age may lead to physiological changes in the respiratory system, poor airway clearance and reduced lung reserve, and defense barrier mechanisms[33].

In terms of blood biochemical findings, we also found that glucose, creatinine, and urea were significantly higher in older patients with COVID-19 infection than in adults or children. In contrast, studies have shown that there was no significant difference in serum creatinine when the elderly were compared with young and middle-aged patients [13]. We showed that 40% of the patients had various comorbid conditions, which might have explained the increased creatinine and glucose levels in the older adults in this study. Likewise, a recent study indicated that COVID-19 patients with severe conditions had high blood urea and creatinine levels, and that their lymphocyte counts continued to decrease [25]. In our results, the LDH level was elevated in all age groups, and indicated an adverse clinical outcome. Elevated LDH has been found to be an independent risk factor for severe/critical COVID-19 in patients [7]. Research data suggest that SARS-CoV-2 may damage liver and myocardium tissues, lead to elevated levels of AST, CK, and myoglobin to various degrees, and increased troponin in critical patients [9]. Tsui et al. showed that LDH levels reflect tissue necrosis related to immune hyperactivity in SARS, and thus relate to poor outcome [34]. Therefore, the monitoring of LDH levels and other cardiac and liver enzymes could help predict severe COVID-19 in patients [12].

Determining the levels of D-dimer, CRP, ESR, and other inflammatory biomarkers is essential to detecting bacterial infection in the lungs, and may help preliminarily evaluate patients’ immune status [26]. We found that most COVID-19 patients had elevated ESR rates, and over 40% of the patients presented increased CRP and D-dimer levels. Likewise, studies have documented that an increased D-dimer concentration is a common feature of COVID-19 infections, especially for severe patients [9,25,26]. However, the elderly are more likely to develop a severe disease [21]. Our findings revealed significantly higher levels of ESR and D-dimer in the elderly than in the adult group, while there were no significant differences in CRP levels between the two groups. A recent study found that the CRP level in elderly patients was significantly higher than that in young and middle-aged groups [13].

Our findings revealed that many patients infected by SARS-CoV-2 were older men and had chronic underlying diseases. We found that diabetes mellitus, renal failure and hypertension were present in 27.0%, 13.9% and 10% of the cases, respectively. This is in agreement with a previous report, in which COVID-19 patients were reported to have comorbid conditions [4,19]. In addition, research data show that patients ≥60 years old are at higher risk than children, who are less likely to have an infection, or who show mild to asymptomatic infection [28]. The increased risk of developing severe COVID-19 complications in older people with comorbid conditions have been well documented in the literature [19]. In multivariate analysis of 487 COVID-19 patients including 49 severe cases, Chi et al noted that elder age (OR 1.06 [95% CI 1.03–1.08], p< 0.001), male (OR 3.68 [95% CI 1.75–7.75], p= 0.001), and presence of hypertension (OR 2.71 [95% CI 1.32–5.59], P=0.007) are independently associated with severe disease at admission [4]. Chen et al. [19]suggested that 2019-nCoV is more likely to infect older adult men with chronic comorbidities as a result of the weaker immune functions of these patients [19]. A study indicated that COVID-19 infection progresses rapidly to acute respiratory distress syndrome, septic shock, metabolic acidosis, coagulation dysfunction, and leads to death in older patients and those with comorbid conditions[25]. Therefore, identifying host risk factors associated with severe COVID-19 infections may help in designing specific strategies to prevent and treat the disease[4].

We found that 24% of the patients were admitted to the ICU. This proportion was much higher than the 4.7% reported in a multi-center retrospective study that included 1519 cases from all regions of Saudi Arabia[8]. The mortality rate of COVID-19 in our study was 8.8%, which was higher than the 3.4 % reported in the literature [25]. The overall fatality rate of the disease as found by others was in the range of 3% to 14%[35]. The high mortality rate and ICU admission observed in this study could be explained by the fact that the study selected only confirmed cases that were admitted to our hospital. Therefore, a further survey with a larger sample size could show some differences. In comparing the death rate of COVID-19 with other coronaviruses, the available data indicate that COVID-19 infection results in a lower mortality rate than that reported for SARS (9.60%) and MERS (34.4%)[36]. Therefore, extensive studies of the pathogenic and virulence mechanisms of SARS-CoV, SARS-CoV-2, and MERS□CoV are needed to explain these variations.

We found that the mortality rate of COVID-19 infection was quite high among the elderly and those admitted to the ICU, which resembled previous findings[13,29] However, patients in the ICU were more likely to receive mechanical ventilation, surgical interventions, and prolonged treatment, which might have increased the death rate[29]. In addition, the study also revealed that hypertension, renal failure, and cardiovascular disease increased the risk of death in COVID-19 cases. This agrees with previous reports, in which hypertension, coronary heart disease, and diabetes were found to be risk factors that resulted in death[12,27,29,36]. Research evidence indicates that the elderly and those with chronic underlying diseases develop severe and fatal respiratory failure because of alveolar damage from the virus [17,19,21].

### Limitations

Our study has several limitations: First, the sample size was relatively small, especially for the children’s group, and may not fully reflect the characteristics of the disease for this group. Therefore, a large sample size could give a more comprehensive understanding of 2019-nCoV. Second, the study findings might have been biased by reporting only confirmed cases in a single hospital center. Further studies that include suspected and undiagnosed cases may show some differences. Third, we statistically analyzed the laboratory findings based on a comparison of means, median, and proportions between different age groups that were not subdivided into groups of patients with individual comorbid conditions. Finally, the study assessed the epidemiological, laboratory, and clinical characteristics of COVID-19 on admission of the patients; more detailed information from other laboratory tests and clinical outcomes were unavailable at the time of analysis.

## Conclusions

In summary, this study revealed the epidemiological, clinical, and laboratory findings of patients of different age groups confirmed to have COVID-19. On admission, patients were most likely to present with mild symptoms of fever, cough, shortness of breath, and chest pain. Increased LDH, CRP, ESR, and D-dimer levels were common laboratory findings on patient admission. However, glucose, CK-MB, LDH, bilirubin, D-dimer, and ESR levels were significantly higher in the elderly than in adult patients. We found an increased disease mortality rate, which may be attributed to the high percentage of patients with various comorbidities. This study revealed that older age, ICU admission, cardiovascular disease, hypertension, and renal failure were risk factors for death. Increased levels of leucocytes, potassium, and urea, and decreased hemoglobin concentration were frequent laboratory findings associated with death. The common symptoms found in this study may help in identifying potential severe COVID-19 patients. Moreover, some routine laboratory findings could help assess disease progression and manage patients who could develop critical conditions and be at risk of mortality. These findings should be confirmed in future studies coupled with testing other suitable clinical and laboratory indicators for evaluating disease severity and outcomes.

## Data Availability

The data supporting the findings of this study are available within the manuscript

## Acknowledgments

The authors extend their appreciation to the Deanship of Scientific Research at University of Bisha, Saudi Arabia, for funding this work through the COVID-19 Initiative Project under Grant Number (UB-COVID-13-1441).

